## Supplemental Table 1 for "Prospective Cohort Study of Surgical Site Infections Following Single Dose Antibiotic Prophylaxis in Caesarean Section at a Tertiary Care Teaching Hospital in Medchal, India"

Supplementary Tables

Table S1: Clinical profile of the study participants based on developing the SSI

| Variable | With SSI | Without SSI | P-value |
| --- | --- | --- | --- |
|  | n=92 (4.57) | n=1924 (95.43) |  |
| Age (in years) Mean±SD | 23.32 ± 2.75 | 24.11 ± 3.25 | 0.0209 |
| **BMI** | | | |
| Underweight | 6 (6.52) | 221 (11.49) | 0.013 |
| Normal | 22 (24. 91) | 582 (30.27) |  |
| Overweight | 11 (11.96) | 210 (10.92) |  |
| Obesity | 25 (27.17) | 279 (14.51) |  |
| Missing | 28 (30.43) | 631(32.81) |  |
| **Booking status** | | | |
| Booked | 83 (90.22) | 1720(89.44) | 0.813 |
| Unbooked | 9 (9.78) | 203 (10.56) |  |
| **No. of prenatal visits** | | | |
| <4 | 9 (9.78) | 181 (9.41) | 0.980 |
| 4-8 | 37 (40.22) | 801 (41.65) |  |
| >8 | 37 (40.22) | 738 (38.38) |  |
| Missing | 9 (9.78) | 203 (10.56) |  |
| **Anaemia (< 11gm%)** | | | |
| Yes | 9 (9.78) | 172 (8.94) | 0.784 |
| No | 83 (90.22) | 1751 (91.06) |  |
| **Hypertension** | | | |
| Yes | 14 (15.22) | 287 (14.92) | 0. 939 |
| No | 78 (84.78) | 1636 (85.08) |  |
| **Hypothyroidism** | | | |
| Yes | 10 (10.87) | 258 (13.42) | 0.482 |
| No | 82 (89.13) | 1665 (86.58) |  |
| **Hospital stay in days Mean±SD** | 11.80 ± 3.65 | 9.29 ± 3.11 | <0.001* |

Table S2: Obstetric related factors of the study participants based on developing the SSI

| Variable | With SSI | Without SSI | P-value |
| --- | --- | --- | --- |
|  | n=92 (4.57) | n=1923 (95.43) |  |
| **Gestational age at delivery (weeks) Mean±SD** | 37.7 ± 1.47 | 37.52 ± 1.66 | 0.1215 |
| **Parity** | | | |
| Primi | 51 (55.43) | 690 (35.88) | <0.001* |
| Multi | 41 (44.57) | 1233 (64.12) |  |
| **Gestational Diabetes** | | | |
| Yes | 3 (3.26) | 78 (4.06) | 0.704 |
| No | 89 (96.74) | 1845 (95.94) |  |
| **PROM** | | | |
| Artificial Rupture of membranes | 57 (61. 96) | 1388 (72.18) | 0.183 |
| Spontaneous rupture of membranes (SROM) <6 h | 25 (27.17) | 355 (18.46) |  |
| SROM (6-12h) | 4 (4.52) | 54 (2.81) |  |
| SROM (>12h) | 6 (6.52) | 113(5.88) |  |
| Not recorded | 0 (0.00) | 13 (0.68) |  |
| **Duration of labor (hours)** | | | |
| <6 | 29 (31.52) | 613 (31.88) | 0.012 |
| 6-12 | 24 (26.09) | 357 (18.56) |  |
| >12 | 13 (14.13) | 126 (6.50) |  |
| Not in labor | 26 (28.26) | 823(42.80) |  |
| Not recorded | 0 (0.00) | 5 (0.26) |  |
| **Chorioamnionitis** | | | |
| Yes | 0 | 2(0.10) | 0.757 |
| No | 92 (100) | 1921 (99.90) |  |
| **Steroids given** | | | |
| Yes | 29 (31.52) | 611 (31.77) | 0.960 |
| No | 63 (68.48) | 1312 (68.23) |  |
| **Previous surgery** | | | |
| Caesarean section | 34 (36.96) | 1077 (56.01) | <0.001 |
| Laparotomy | 0 | 5 (0.26) | - |
| Caesarean section and laparotomy | 0 | 13 (0.68) | - |
| No | 58 (63.04) | 827 (43.01) | - |
| Not recorded | 0 | 1 (0.05) | - |

Table S3: Anaesthesia and operative related factors of the study participants based on developing the SSI

| Variable | With SSI | Without SSI | P-value |
| --- | --- | --- | --- |
|  | n=92 (4.57) | n=1923 (95.43) |  |
| **Time interval between hair removal and surgery (hours)** | | | |
| <2 | 15 (16.48) | 255(13.25) | 0.335 |
| >2 | 74(81.32) | 1664(86.49) |  |
| **Type of LSCS** | | | |
| Elective | 22 (23. 91) | 767 (39.89) | 0.002* |
| Emergency | 70 (76.09) | 1156 (60.11) |  |
| **Type of skin disinfectant used** | | | |
| Povidone iodine | 0 (0.00) | 7 (0.36) | 0.127 |
| Chlorhexidine | 1 (1.09) | 3 (0.16) |  |
| 70% alcohol | 1 (1.09) | 3 (0.16) |  |
| Povidone iodine + chlorhexidine | 0 (0.00) | 5(0.26) |  |
| Povidone + 70% alcohol | 90 (97.82) | 1898 (98.70) |  |
| Povidone iodine+Chlorhexidine+ 70% alcohol | 0 (0.00) | 7 (0.36) |  |
| **Type of anaesthesia used** | | | |
| Spinal | 92 (100.00) | 1916 (99.64) | 0.845 |
| Epidural | 0 (0.00) | 2 (0.10) |  |
| General | 0(0.00) | 5 (0.26) |  |
| **ASA score** | | | |
| 1 | 89 (96.74) | 1874 (97.45) | 0.880 |
| 2 | 3 (3.26) | 48 (2.50) |  |
| 3 | 0(0.00) | 1 (0.05) |  |
| **Type of incision** | | | |
| Pfannensteil | 91 (98.91) | 1904 (99.01) | 0.926 |
| Subumbiical midline | 1 (1.09) | 19 (0.99) |  |
| **No. of persons in OT** | | | |
| <5 | 7 (7.61) | 43 (2.24) | 0.011* |
| 5-10 | 57 (61. 96) | 1186 (61.67) |  |
| 11-15 | 27 (29.35) | 657 (34.17) |  |
| >15 | 1 (1.09) | 37 (1.92) |  |
| **Duration of surgery in minutes (Mean±SD)** | 45.12 ± 10.24 | 47.29 ± 12.72 | 0.1064 |
| **Exteriorization of uterus** | | | |
| Yes | 16 (17.39) | 398 (20.70) | 0.443 |
| No | 76 (82.61) | 1525 (79.30) |  |
| **Delivery by forceps** | | | |
| Yes | 27 (29.35) | 637 (33.13) | 0.451 |
| No | 65 (70.65) | 1286 (66.87) |  |
| **Use of cautery in subcutaneous tissue** | | | |
| Yes | 2(2.17) | 12 (0.62) | 0.080 |
| No | 90 (97.83) | 1911 (99.38) |  |
| **Placental removal** | | | |
| Spontaneous | 89 (96.74) | 1891 (98.34) | 0.252 |
| Manual | 3 (3.26) | 32 (1.66) |  |
| **Type of skin closure** | | | |
| Mattress | 2 (2.17) | 42(2.18) | 0.995 |
| Subcuticular | 90 (97.83) | 1881 (97.82) |  |
| **Suture material used** | | | |
| Sutupack | 4 (4.39) | 207 (10.76) | 0.03* |
| Silk | 84 (92.31) | 1632 (84.82) |  |
| Sutupack + Silk | 2(2.20) | 78 (4.05) |  |
| Monocryl | 0 | 5 (0.26) |  |
| Others | 1 (1.10) | 2(0.11) |  |
| **Prophylactic antibiotic administered** | | | |
| Yes | 88 (95.65) | 1897 (98.65) | 0.020 |
| No | 4 (4.39) | 26 (1.35) |  |
| **Duration of antibiotic given (Mean±SD)** | 10.70 ± 5.49 | 10.73 ± 5.50 | 0.564 |
| **Type of prophylactic antibiotic given** | | | |
| Inj Cefazolin | 45 (48.91) | 864(44.93) | 0.456 |
| Inj.Ampicillin | 24 (26.09) | 517 (26.89) | 0.865 |
| Inj.Taxim | 19 (20.65) | 513 (26.68) | 0.165 |
| Others | 0 | 3 (0.16) | - |
| Not given | 4(4.35) | 26 (1.35) | - |
| **Intraoperative blood transfusion given** | | | |
| Yes | 1 (1.09) | 32 (1.66) | 0.670 |
| No | 91 (98.91) | 1891 (98.34) |  |
| **UTI developed after surgery** | | | |
| Yes | 2 (2.17) | 3 (0.16) | <0.001* |
| No | 90 (97.83) | 1920 (99.84) |  |
